# Revolutionary High-Performance Soluble Collagen-Based Surgical Sutures: The Next Generation Tissue Healing Technology

**DOI:** 10.1101/2025.05.19.25325989

**Authors:** Xiaoming He, Zhaohui Luo, Shenghua He, Wenbin Jin

## Abstract

This study presents an innovative absorbable surgical suture designed to overcome the limitations of conventional absorbable sutures. Traditional collagen-based catgut sutures elicit immune reactions and undergo uncontrolled degradation. The currently prevalent absorbable synthetic polymer sutures exhibit high strength, but their degraded byproducts can damage tissue cells. Consequently, they are not suitable for certain surgeries, particularly in sensitive areas such as neuro-dense sites.

Through advanced material modification and processing techniques, we have developed a novel absorbable suture based on soluble collagen. This suture maintains exceptional biocompatibility and low antigenicity, while simultaneously enhancing mechanical strength and extending its degradation cycle. Animal experiment and preliminary human application demonstrate reduced postoperative inflammation, improved wound healing, and minimized scarring. Notably, the collagen suture exhibits higher mechanical strength in vivo compared to control synthetic polymer sutures. Furthermore, the suture’s degradation is synchronized with wound healing, and no immune rejection or adverse events were observed.

This suture offers significant advantages in sensitive tissue applications, such as ophthalmology and neurosurgery, eliminating the need for removal procedures. It represents a paradigm shift in suture technology, providing a revolutionary approach to precise medical interventions. This material technology has the potential to transform global surgical practices.

## Introduction

Surgical sutures, fundamental tools in medical practice, have evolved over thousands of years, ancient Egypt’s linen threads and India’s cotton threads have historical significance [Babel and Thomas, 2024].

Collagen-based catgut sutures were introduced in the 18th century, but their modern use is limited by issues such as immune rejection and unpredictable degradation [Li et al., 2018]. Suture materials have undergone significant transformations, with the mid-20th century witnessing a revolutionary leap with the introduction of synthetic absorbable sutures like PCL, PLA, PGA and their co-polymer [Dennis et al., 2016; Muffly et al., 2011, Pillai and Sharma, 2010]. Despite these developments, current absorbable synthetic sutures still present challenges such as foreign body reaction rates (up to 18.7%), tissue irritation, and inconsistent degradation in specific environments [Hochberg et al., 2009]. These issues are particularly critical in neurosurgery and ophthalmology, underscoring the urgent need for sutures optimized for sensitive tissues [Shai et al., 2007]. The quest for “ideal suture materials” continues, aiming for predictable mechanical properties, controlled degradation, minimal tissue reactions, and true biocompatibility [Ethicon Inc., 2019].

Soluble collagen, the body’s most abundant structural protein, possesses low antigenicity and strong tissue affinity, making it theoretically ideal for surgical sutures [Parenteau-Bareil et al., 2010]. However, soluble collagen exhibits low mechanical strength, strong hydrophilicity, and high solubility, and degrades rapidly in vivo, restricting its further medical application. Attempts to enhance collagen’s mechanical strength through chemical cross-linking improved durability but did not yield substantial achievements, while biotoxicity and tissue inflammation increased [Dasgupta et al., 2021]. Genetically engineered recombinant collagen reduced antigenicity but encountered production challenges, hindering its large-scale medical application [Fertala, 2020]. Despite these advancements, achieving the optimal collagen-based suture remains a formidable challenge.

Building upon prior research, our team has developed a novel absorbable surgical suture based on soluble collagen. This advancement was achieved through innovative processing techniques and multi-level cross-linking strategies. This advancement significantly improves mechanical strength, degradation control, and biocompatibility. Key innovations include the elimination of major immunogenic epitopes in collagen, the improvement of material strength and its molecular structure, the optimized cross-linking regulating degradation while maintaining low antigenicity, and the reduction of postoperative inflammation and irritation. These features make the sutures ideal for sensitive applications in ophthalmology and neurosurgery, offering unprecedented surgical options for high-risk tissues[Sullivan et al., 2023]

In comparison to Vicryl rapid sutures (Ethicon Inc.), collagen sutures demonstrate promising results in both animal and preliminary human trials. These sutures exhibit reduced postoperative inflammation, enhanced wound healing, and minimized scarring. The medical advantages of collagen sutures open up new avenues for biomedical material development.

With over 234 million surgeries performed annually, accounting for 15% of sensitive tissue procedures [Thomas, 2008], this innovation presents a transformative potential for patient outcomes, particularly in resource-constrained settings. By eliminating the necessity for suture removal, it can significantly reduce complications and associated costs.

This research investigates the identified challenges and limitations of existing absorbable sutures. It synergistically combines the research findings of our developed collagen sutures to assess their application value and delineate future development directions.

## Materials and Methods

### Fabrication of Absorbable Collagen Sutures

Collagen threads were fabricated using a medical-grade, soluble collagen derived from pig skin with a molecular weight of approximately 300,000 Dalton. After adjusting pH and temperature, this solid collagen was dissolved to form a solution with a concentration of at least 3% (a low concentration is suitable for collagen sutures with small diameters). Subsequently, a small amount of chemicals such as hyaluronic acid, proteoglycan, Glucosamine, chondroitin sulfate, protein polysaccharide, were appropriately incorporated into the collagen solution based on the intended application (strength or affinity).

After mixing, filtration, and defoaming, the collagen solution underwent a uniform treatment to eliminate gel blocks and bubbles. Subsequently, it was compressed and extruded from a spinneret (e.g., a syringe needle) into a coagulation bath (e.g., isopropanol or alcohol). This process resulted in the formation of collagen threads.

The collagen threads were subsequently immersed in a cross-linking solution (e.g. glutaraldehyde, carbodiimide, urea-formaldehyde resin, epoxy resin, genipin) for a predetermined duration (its in vitro degradation time increases with the increase of cross-linking time). Upon removal from the cross-linking solution, the collagen threads were fully dried in clean air. This process yielded absorbable collagen sutures. Various diameters of sutures can be manufactured in this manner. Different colors of collagen sutures (threads) can be obtained by adding various ingredients to the collagen solution and employing different cross-linking methods. Two colors were fabricated in the experiment: transparent and black. The samples of the collagen sutures with two colors and eight diameters are presented in Figure 1.

**Figure 1.**
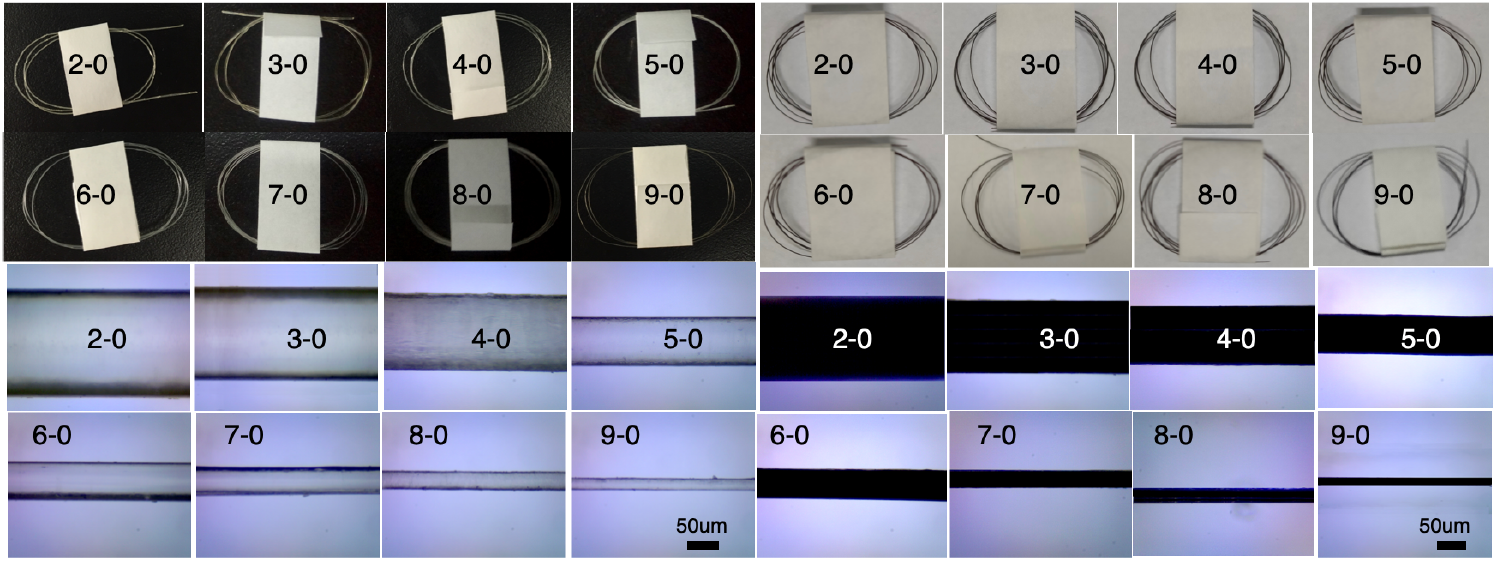
Transparent and black absorbable collagen sutures with various diameters. Left: Transparent sutures; Right: Black sutures; Lower: Microscope observations. Note: Suture 2-0 has a large diameter, while 9-0 has a smaller diameter.

### Evaluation of the Properties of Collagen Sutures

The properties of absorbable collagen sutures are officially evaluated by the Guangdong FDA, which is affiliated with the China Food and Drug Administration (CFDA). The evaluation categories encompass the mechanical strength and biological properties of collagen sutures. The evaluation of biological properties includes cytotoxicity tests, intradermal stimulation tests, hemolysis tests, systemic toxicity tests, sensitization tests, pyrogen tests, implantation tests, and genotoxicity tests.

### Skin suture experiment (Suture Specification: 5-0)

Animal Model: eighteen male New Zealand rabbits weighing 2.0-2.7 kg, were utilized.

Vicryl rapid sutures (Specification: 5-0, W9915, Ethicon, Johnson & Johnson Medical Equipment Co., Ltd.) were utilized as a control, which are a copolymer of glycolide and lactide 910.

All animal procedures were meticulously conducted in accordance with the applicable laws, regulations, and institutional guidelines governing the use and care of laboratory animals. Animal welfare and ethical considerations were strictly adhered to throughout the study.

Sixteen hours prior to the surgical procedure, the rabbits were fasted. The surgical area was meticulously shaved and thoroughly disinfected with iodine. The animals were anesthetized by a 10% hydrated chloraldehyde peritoneal injection (1mL/kg), concurrently administered with a fast-sleep-new II intramuscular injection (0.3mL/kg). Immediately following the operation, penicillin sodium (200,000 units/rabbit) was administered intramuscularly and continued to be administered within the subsequent three days.

Four 3 cm incisions were made on the back of a rabbit. These incisions were positioned on both sides of the rabbit’s back, with two incisions on each side. The upper left incision was closed with a test suture (collagen suture), while the upper right incision served as the control suture (Vicryl rapid suture). The right lower incision was embedded with the control suture, while the left lower incision was embedded with the test suture. Following the incision of the skin, blood vessel forceps were utilized to retract the skin inward, forming a 2 cm deep capsule. A 10 cm long suture was weighed and inserted into the capsule, followed by suturing with standard 4-0 silk sutures.

Following implantation, the animal was individually fed per cage, with access to water provided freely. On-demand feeding was implemented, and daily observations and recordings were conducted to monitor animal activities, wound healing, incision inflammation, and infection. Weekly weighings were conducted throughout the experiment.

### Mechanical Strength of Skin Anastomosis

Skin incision and their surrounding skin were excised at 3, 7, and 18 days post-operative, leaving approximately 4 cm of skin on both sides of the incision. The mechanical strength of the skin anastomosis was evaluated using a universal testing machine.

### Pathological Examination

Following evaluation of the mechanical strength of the skin anastomosis, the skin was fixed with neutral formaldehyde, embedded in paraffin, and sectioned. Subsequently, it was stained with Hematoxylin and eosin (HE) and Masson stains to assess the pathological alterations of the skin anastomosis, including inflammation and alterations in collagen fibers.

### Suture Degradation in vivo

3, 7, and 18 days after the surgery operation, the embedded sutures were removed, and the attached tissue and mucus were gently wiped with a sodium chloride solution. The sutures were subsequently dried in a vacuum drying oven for seven days, and their weights were recorded. The degradation rate of the sutures was calculated using the following formula:

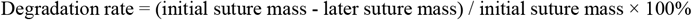

### Suture Mechanical Strength after Operation

The mechanical strength of the suture was determined using a universal testing machine (AX-400), and its diameter was also measured. The mechanical strength of the suture was subsequently calculated using the following formula:

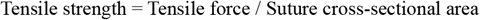

### Preliminary human application

The study encompassed a single human case in the cosmetic plastic surgery department of Tongji Hospital, which is affiliated with Tongji Medical College of Huazhong University of Science and Technology. The surgical procedure entails the removal of the prosthesis that occupies the breast and the subsequent suturing of the incision utilizing collagen sutures. This observation was approved by Ethics Committee of Huazhong University of Science and Technology, under reference number [2019EA254], and informed consent was obtained from the patient regarding the experimental nature of the procedure.

#### Statistics

All data were presented as Mean **±** Standard Deviation (SD) and analyzed using Microsoft Excel software. A t-test was employed to assess the disparity between the two groups. A p-value less than 0.05 indicates a statistically significant difference.

## Results

Figure 1 shows transparent and black absorbable collagen sutures with various diameters. All sutures are single strands. Sutures with large diameters (such as 2-0) are rigid and difficult to knot. They can be fabricated (weaved or twisted) by collagen sutures with small diameters (such as 9-0).

The collagen suture exhibits remarkable mechanical strength, surpassing the standards of surgical sutures, and possesses exceptional biological properties, as officially recognized by the CFDA (No. MZ17010224, No. WT17080974, No. WT19010144, No. WT19010145). Details of biological properties of the absorbable collagen suture were evaluated and showed in Table 1:

**Table 1.** The biological properties of the absorbable collagen suture.

| Test Items | National Standard | Test results |
| --- | --- | --- |
| 1 Cytotoxicity | <class 1 | Class 0 |
| 2 Intracutaneous test | No stimulation | No stimulation |
| 3 Hemolysis | Hemolysis rate<5% | Hemolysis rate 0.6% |
| 4 Systemic Toxicity | No toxicity | No toxicity |
| 5 Sensitization | No reaction | No reaction |
| 6 Pyrogen | No Pyrogen | No Pyrogen |
| 7 Implantation | No inflammatory reaction | No inflammatory reaction |
| 8 Genotoxicity |  |  |
| Chromosome aberration | No toxicity | No toxicity |
| Ames | No toxicity | No toxicity |
| Mouse lymphoma | No toxicity | No toxicity |

Table 1 shows that the collagen suture’s biological properties match international standards for absorbable surgical sutures. This suggests it has excellent implant properties, potentially leading to superior outcomes than conventional sutures.

### Preliminary human application

**Figure 2** illustrates suturing skin wounds using black (top images) and transparent (middle & bottom images) collagen sutures. The top images depict black sutures in rabbits, the middle images depict transparent sutures in rabbits, and the bottom image depicts a transparent suture in a patient (woman). Although it may be challenging to distinguish the transparent surgical sutures from the rabbit hair, it is evident that the collagen suture has completely degraded by day 10 (top and middle), and back wound healing was perfect at day 10. There appears to be no discernible distinction in the healing process of skin wounds between black and transparent collagen sutures. The lower figures illustrate suturing an incision wound after removing a prosthetic from a woman’s breast. They demonstrate that the suture began to degrade at day 6, with two visible collagen suture ends (red squares) while the other suture parts remain within the skin (yellow circle indicates needle holes). However, by day 9, the suture has almost completely degraded, leaving only one suture end (red square), and the skin has healed remarkably well at that point. From these images of wound healing in animals and humans, it becomes evident that collagen suture degradation is closely synchronized with the wound healing process.

The patient and the doctor are satisfied with the absorbable collagen sutures, as non-absorbable sutures, such as nylon, silk, and polypropylene sutures, commonly used in plastic surgery, are typically removed approximately 7-10 days after surgery. This type of absorbable collagen suture facilitates a smooth healing process with minimal inflammation and discomfort, and it does not require removal.

**Figure 2.**
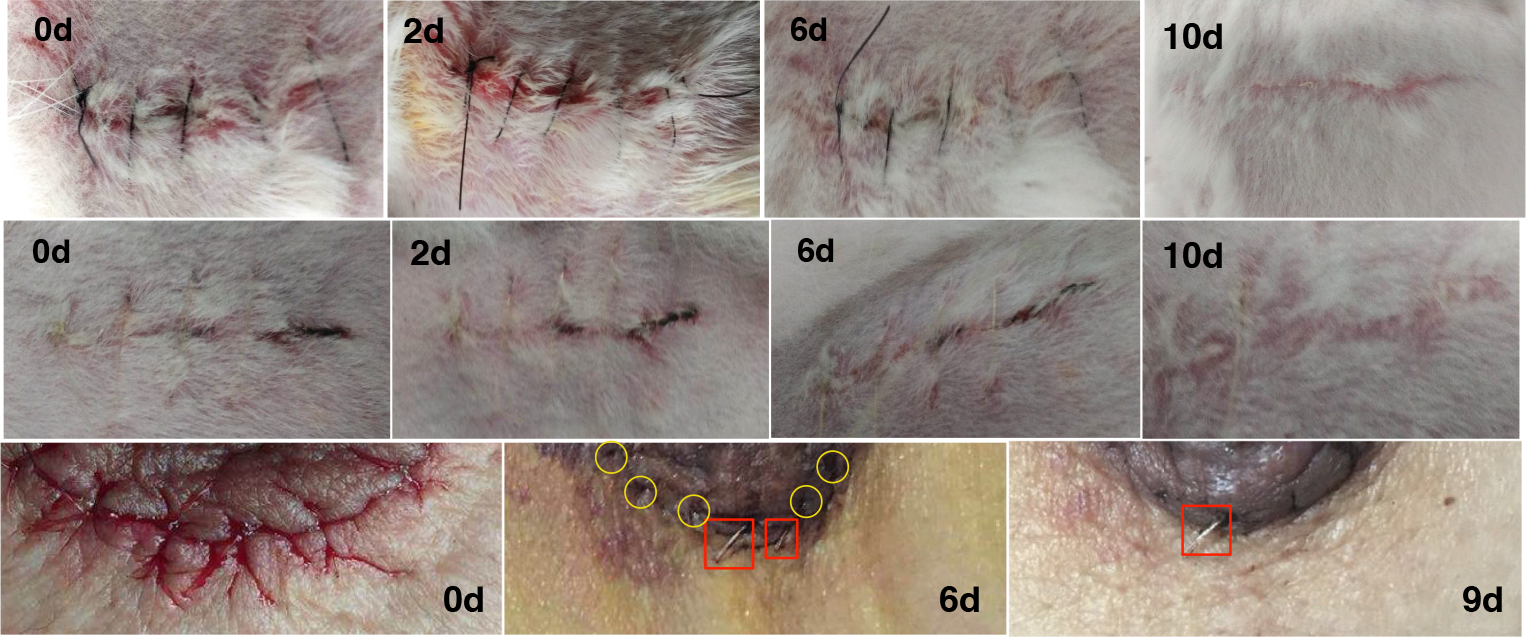
Skin Wounds After Suturing Using Collagen Sutures Top: Black sutures in a rabbit’s back; Middle: Transparent sutures in a rabbit’s back; Bottom: Transparent sutures in a patient’s breast.

### Data of animal experiments

#### Animal Experiment Observation

In each group, the rabbits underwent suturing with collagen suture (test) and Vicryl rapid suture (control). Notably, no substantial weight loss was observed in any group following the surgical procedure. Additionally, no wound disintegration, reinjury, or infection was detected postoperatively. Both the test and control sides exhibited satisfactory wound healing.

#### Pathological examination

At 3, 7, and 18 days post-suturing operation, the skin wounds on the test and control sides healed remarkably well. Hematoxylin and eosin (HE) and Masson stain examinations of the skin anastomosis were conducted, and the results are presented in Figure 3. A small amount of fibroblasts and a small amount of inflammatory cell infiltration were observed around the wound healing site of both groups. It appears that test groups utilizing collagen sutures exhibit more aesthetically pleasing outcomes, while there is no discernible distinction between the control and test groups.

**Figure 3.**
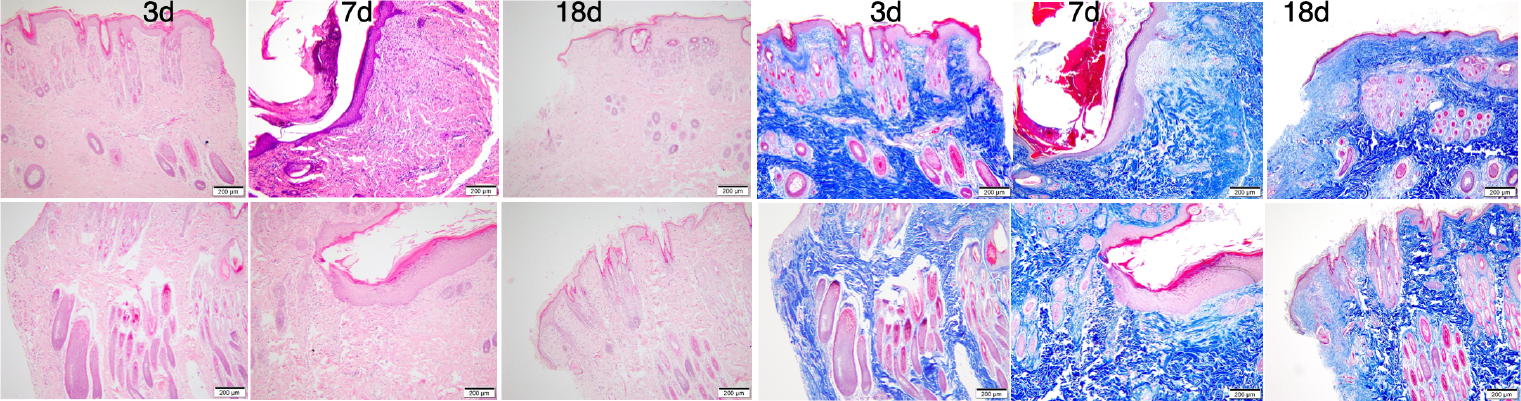
HE stain (left pink) and Masson stain (right blue) of skin anastomoses at 3, 7,18 days post-suturing operation using collagen suture (top) and control suture (bottom).

#### Mechanical Strength of Rabbit Skin

The mechanical strength of rabbit healed skin was measured. The results demonstrated a correlation between the skin’s strength and the duration of healing. At 3, 7, and 18 days post-operation, the skin strength of the test side exhibited a increase compared to the control side, as evidenced in Figure 4.

**Figure 4.**
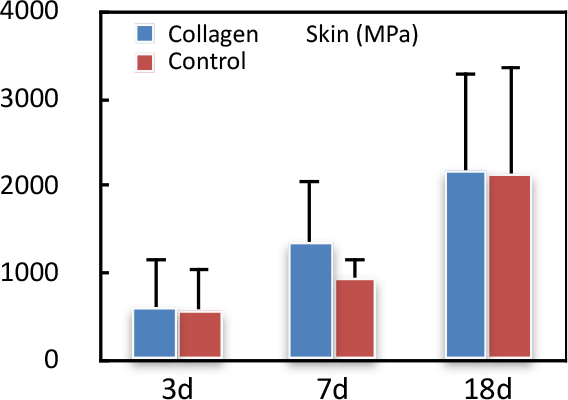
Strength of rabbits’ healed skin at 3d, 7d, 18d (n = 6)

#### Degradation of suture

At 3, 7, and 18 days post-subcutaneous implantation, the sutures from the test samples and control samples were excised and weighed. Due to water absorption and cell adhesion, the weight was higher than that before implantation (day 0). The cell adhesion was removed, and the sutures were dried for 7 days. Subsequently, the degradation rate of the sutures was calculated and showed in Fig.5. The results demonstrated a statistically significant (P < 0.05) increase in the degradation rate of the test suture compared to the control suture at both 3 and 7 days post-subcutaneous implantation. This observation clearly demonstrates that collagen sutures undergo a more rapid degradation compared to control sutures. However, at 18 days of implantation, the degradation rate of the test suture did not exhibited a statistically significant higher rate in comparison to the control suture.

**Figure 5.**
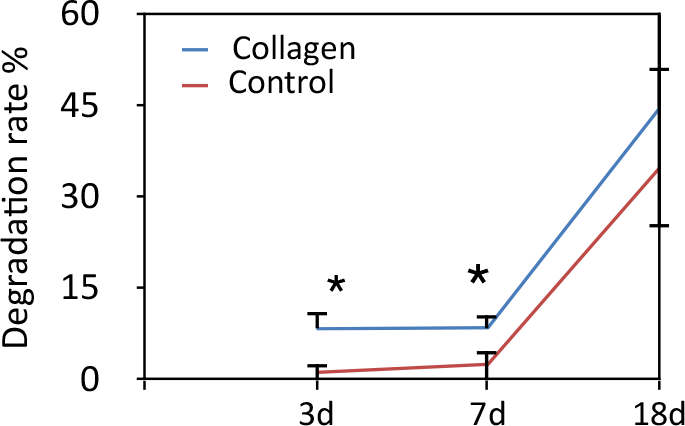
Degradation rate of test and control sutures at different time points (n = 6). Note: * P < 0.05.

#### Tensile Strength of Sutures

After subcutaneous implantation for 3 and 7 days, the sutures were removed and dried, respectively. The tensile strength of the sutures at two time points was measured and recorded in Figure 6. Statistically, the strength (MPa) and force (N) of collagen sutures were significantly higher than those of the control sutures (P < 0.05).

**Figure 6.**
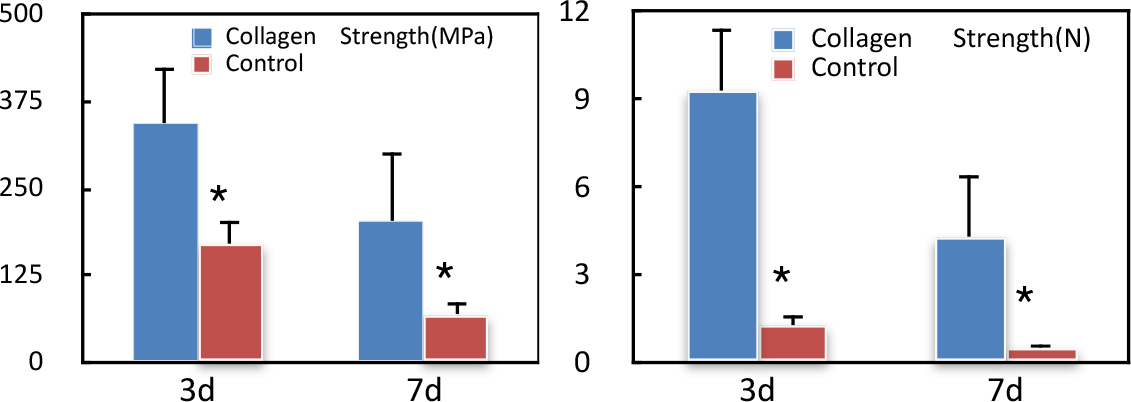
Suture tensile strength at different time points (n = 6). * P < 0.05. Note: At 18 days, due to the sufficient degradation, the sutures fractured into broken parts, rendering their strength unmeasurable.

## Discussion

This research has successfully developed an innovative surgical suture based on soluble collagen and demonstrated its unique advantages through animal test and preliminary human application. In a 3cm wound suturing experiment on rabbit backs, compared with the commercial Vicryl rapid suture (Johnson & Johnson, w9915, Ethicon), the collagen suture showed superior wound healing effect and quality: healed wounds showed more aesthetic appearance with reduced scarring, consistent with the mechanism of collagen biomaterials promoting tissue reconstruction [Rezvani et al., 2021]. Tensile strength of healed wounds showed that samples using the collagen suture had breaking strengths higher than the control group, supporting the positive impact of collagen materials on tissue repair quality[Chattopadhyay and Raines, 2014].

Collagen sutures demonstrate synchronization of degradation with wound healing. Healed skin exhibits enhanced strength in the collagen suture group compared to the control group. This indicates that cells degrade collagen as they acquire growing nutrition to repair the wound. Once the wound heals well, the collagen suture was assimilated and disappeared. In other words, the suture’s degradation rate closely aligns with the wound healing process, outperforming the control group’s, which is highly consistent with concept that “ideal biomaterials should degrade synchronously with tissue reconstruction” [Bonferoni et al., 2021]. Our experimental data results proved that medical devices based on biologically homologous materials can better coordinate with human physiological processes.

In preliminary human application, collagen sutures have achieved satisfactory results in preliminary cosmetic plastic surgery. These sutures have demonstrated positive outcomes in both surgeon operation evaluations and patient satisfaction surveys.

The observed improvement in healing quality may be based on reduced inflammatory response, inflammatory responses around collagen materials were lower than around synthetic materials. Reduced inflammation helps decrease scar formation and improve healing aesthetics. Collagen molecules possess remarkable compatibility, low antigene content, and high nutrient value. These attributes facilitate the stimulation of the migration and proliferation of fibroblasts and keratinocytes, which are fundamental for the efficient closure of wounds [Mathew-Steiner et al., 2021]. Additionally, collagen sutures may act as biological scaffolds for tissue regeneration, providing an ideal three-dimensional microenvironment for cells’ growth.

### Comparison with Traditional Absorbable Sutures

Absorbable sutures widely used in modern surgery are mainly based on synthetic polymers, such as PGA, PLA, PCL etc. The main advantages of these materials are high mechanical strength and controllable degradation periods, however, multiple studies have shown that these heterologous synthetic materials induce significant foreign body reactions in vivo [Abhari et al. 2017]. Faris et al. (2022) systematically demonstrated that PLGA material degradation products stimulate macrophages to secrete inflammatory factors IL-1β and TNF-α, leading to local inflammatory responses and tissue damage. Li(1999) comparative study found that synthetic polymer suture materials cause local pH decreases during in vivo degradation, creating an acidic tissue environment that further exacerbates inflammatory responses. This aligns with our findings in animal experiments, wherein collagen sutures demonstrated superior outcomes.

Traditional collagen suture materials (such as catgut sutures) have long been limited in application due to their immune reaction and unpredictable degradation rates. Traditionary tissue-based collagen materials release antigenic fragments during in vivo degradation, inducing immune responses [Jin et al., 2024]. In contrast, soluble collagen has very low antigenicity, through specific modifications and processing techniques, we have furthermore reduced the material’s antigenicity, consistent with the finding that collagen’s antigenicity mainly comes from specific amino acid sequence regions [Shoulders et al., 2009]. Mechanical strength of collagen materials was improved through chemical cross-linking but cross-linking agent residues often increased material toxicity [Kirk et al., 2013]. Our technology successfully improved material strength through biocompatible cross-linking methods while maintaining excellent biosafety, paving the way for collagen applications in broader medical fields.

### An interesting phenomenon of suture degradation

A significant finding from this study is that collagen sutures demonstrate relatively stable high strength during the initial 3-7 days of in vivo use, whereas control group synthetic materials rapidly lose strength. Notably, collagen sutures undergo more rapid weight loss (degradation) compared to the control group. This apparent contradiction arises from the distinct degradation characteristics of collagen sutures and synthetic polymer sutures, such as Vicryl rapid, indicating that collagen materials may undergo beneficial structural reorganization in the physiological environment.

The fundamental disparities in their degradation mechanisms contribute to these differences in their properties. Specifically, collagen sutures undergo enzymatic degradation (surface erosion), gradually degrading from the outer surface inward. In contrast, Vicryl rapid (PLGA) undergoes hydrolysis degradation (bulk degradation), affecting the material’s overall structure. Hydrolysis causes internal structure loosening, significantly reducing mechanical strength. Degradation generates acidic byproducts, including lactic and glycolic acids, which can potentially lead to local microenvironment acidification. This acidification may necessitate a prolonged elimination time by the body; for instance, the assimilation of Vicryl rapid suture by the body takes approximately 42 days [Barber et al., 2011; Ethicon Inc., 2019]. In contrast, collagen sutures primarily degrade through enzymatic degradation (surface erosion), allowing for gradual degradation from the surface inward. Cell-secreted collagenases, such as MMP-1, MMP-8, and MMP-13, specifically recognize and degrade collagen [Pittayapruek et al., 2016]. This degradation process is more controllable, preserving structural integrity and mechanical properties. The degradation products are peptides and amino acids, as excellent nutritional substances, which are readily absorbed and utilized by the body. As a natural component of the organism, collagen exhibits superior biocompatibility, facilitating cell adhesion, proliferation, and migration. During the degradation process, new intermolecular cross-linking or recombination may occur, temporarily strengthening the remaining structure [Hematti, 2018]. Additionally, collagen’s enhanced integration with host tissue promotes the growth of tissue cells on the suture and the secretion of new extracellular matrix (ECM)[Brown and Philips, 2007]. That is, collagen suture and Vicryl suture show seemingly contradictory but reasonable performance characteristics due to different degradation mechanisms. The quality loss of collagen sutures may be faster, but the residual part maintains higher strength, which is important for the development of a new generation of biomedical suture materials.

### Current Challenges and Future Directions

Despite the significant achievements of this research, we clearly recognize several key challenges that remain: Further Enhancement of Mechanical Strength. Currently, although the collagen sutures demonstrate unique strength maintenance characteristics in vivo, their initial strength is still lower than some commercial synthetic materials, which may limit their application in high-stress areas. For this challenge, we propose the following research directions:

(1)-Molecular-Level Structural Optimization: Improvements on collagen molecular arrangement will enhance the material’s intrinsic strength by regulating hydrogen bonds and hydrophobic interactions between collagen molecules [Amirrah et al., 2022]. (2)-Composite Reinforcement Strategy: Combining biocompatible materials (such as silk protein nanofibers) with collagen to achieve synergistic reinforcement effects. (3)-Biomimetic Cross-linking Technology: enzyme-catalyzed biomimetic cross-linking method effectively improved material strength, avoiding potential toxicity issues of chemical cross-linking agents [Heck et al., 2013].

### Expanding medical applications

Currently, the collagen sutures have achieved good results in cosmetic plastic surgery, dermatology, but applications in other special fields still need further exploration. For these highly specialized fields, we recommend: (1)Ophthalmic Applications: further improve flexibility, and reduce friction coefficients with eye tissues through surface modification to make them suitable for minimally invasive ophthalmic surgery. (2)Neurosurgical Applications: enhance the suture’s promotion of nerve tissue repair by incorporating neurotrophic factors [Huang et al., 2023]. (3)Specification Diversification: develop a product series with different diameters and degradation periods to meet diverse medical needs.

### Conclusion and Outlook

The absorbable collagen surgical suture developed in this research has demonstrated significant innovation value and medical potential in animal experiments and preliminary human applications. Its excellent tissue compatibility, wound healing promotion effects, and synchronization of degradation with healing represent an important breakthrough in the field of surgical suture materials. Particularly in fields like cosmetic surgery that demand high healing aesthetics, the collagen material shows clear advantages.

Although there is still room for improvement in aspects such as mechanical strength, these challenges also point the way for future research. We believe that through continuous technological innovation and multidisciplinary collaboration, absorbable collagen sutures have the potential to play important roles in broader medical fields, especially achieving breakthroughs in sensitive tissue surgeries such as ophthalmology and neurosurgery, providing patients with higher quality medical experiences.

The collagen materials technology can also be utilized to fabricate tubes, patches, membranes, and other similar structures that can be employed for the repair of various soft tissue defects within the human body.

## Data Availability

All data produced in the present study are available upon reasonable request to the authors

